# An observational study of clinical and health system factors associated with catheter ablation and early ablation treatment for atrial fibrillation in Australia

**DOI:** 10.1101/2021.09.03.21263104

**Authors:** Juan C. Quiroz, David Brieger, Louisa Jorm, Raymond W Sy, Michael O Falster, Blanca Gallego

## Abstract

**Objective:** To investigate clinical and health system factors associated with receiving catheter ablation (CA) for non-valvular atrial fibrillation (AF).

**Study Design and Setting:** We used hospital administrative data linked with death registrations in New South Wales, Australia for patients with a primary diagnosis of AF between 2009 and 2017. We investigated factors associated with receiving CA (using Cox regression) and early ablation (using logistic regression).

**Results:** Cardioversion during index admission (hazard ratio [HR] 1.96; 95% CI 1.75 – 2.19), year of index admission (HR 1.07; 1.07; 95% CI 1.05 – 1.10), private patient status (HR 2.65; 95% CI 2.35 – 2.97), and living in more advantaged areas (HR 1.18; 95% CI 1.13 – 1.22) were associated with a higher likelihood of receiving CA. Private patient status (odds ratio [OR] 2.04; 95% CI 1.59 – 2.61) and a history of cardioversion (OR 1.25; 95% CI 1.0 – 1.57) and diabetes (OR 1.6; 95% CI 1.06 – 2.41) were associated with receiving early ablation.

**Conclusion:** Beyond clinical factors, private patients are more likely to receive CA and earlier ablation than their public counterparts. Whether the earlier access to ablation procedures in private patients is leading to differences in outcomes among patients with atrial fibrillation remains to be explored.

What is new?

- Key findings
  - Private patient status had the strongest effect on the likelihood and odds of receiving catheter ablation and early ablation compared to other clinical and health system factors
  - Cardioversion during index admission, year of index admission, and living in more advantaged areas were associated with a higher likelihood of receiving catheter ablation, while older age and a history of congestive heart failure, hypertension, diabetes, and myocardial infarction were associated with a lower likelihood of receiving catheter ablation.
  - Cardioversion during index admission and a history of diabetes were also associated with higher odds of receiving early ablation.
- What it adds
  - Given the growing evidence of the effectiveness of catheter ablation for treating atrial fibrillation, this study demonstrates the associations between clinical and health system factors and receiving catheter ablation and early ablation.
- Implications
  - Clinicians and policymakers should review existing policies to ensure effective procedures for atrial fibrillation are available to the whole population, rather than those who are more advantaged.

## INTRODUCTION

Atrial fibrillation (AF) is the most common cardiac arrhythmia and it is a major driver of cardiovascular hospitalization. One study of AF hospitalizations in Australia from 1993 to 2013 showed national AF hospitalizations increasing 295% over these 21 years [1]. Another study estimated that 5.35% of Australian adults over 55 years of age are affected by AF, with this expected to rise to 6.39% by 2034 [2]. These increasing trends in Australia and across the world [3]–[5] are a cause for concern, as AF is associated with increased risk of stroke, heart failure, and mortality [6], [7].

Guidelines for the diagnosis and management of AF are available [3], [8], [9], including the use of cardioversion, antiarrhythmic medications, and percutaneous catheter ablation (CA). Recent clinical trials have demonstrated the superiority of CA compared to medical therapy across a range of outcomes including maintenance of sinus rhythm, delayed progression to persistent AF, reduced AF-related hospitalization and improved symptoms and quality of life [10]–[12]. These data have also been reflected in recent guidelines recommending CA as preferred therapy in patients who have failed medical therapy (Class I indication) or as an alternative to medical therapy (Class IIa or IIb) [9] and in an increase in the use of CA. In Australia, a 2018 study using data from the Pharmaceutical Benefits Scheme and the Medicare Benefits Schedule found a 48-fold increase (71 to 3480) in the number of CAs from 1997 to 2016 [13].

Randomized controlled trials have shown the efficacy of CA as first-line treatment, with one meta-analysis of six RCTs finding that first-line CA reduced arrhythmic recurrences when compared with anti-arrhythmic drugs [14], [15]. Delivering ablation earlier in treatment of AF may be more effective in maintaining sinus rhythm, has the potential to retard the progression of electro-anatomical changes associated with AF, and may reduce overall health care utilization [14]–[16]. One study in the US identified an increase in the proportion of patients undergoing early ablation from 5% in 2010 to 10.5% in 2016, and the odds of receiving early ablation doubling between 2010 and 2016 [17]. Given the increasing use of CA and the potential benefits of early ablation, it is important to investigate which patients are receiving CA. Prior studies have shown that patients with more comorbidities and higher risk scores are less likely to receive ablation [18] and early ablation [17], and an increasing number of cardioversions are associated with an increased risk for CA [19]. One study of the Outcomes Registry for Better Informed Treatment of Atrial Fibrillation (ORBIT-AF), a registry of AF outpatients in the US, found that patients with a previous CA were younger and more often white, male, and privately insured [10].

To date, no study in Australia has explored patient characteristics associated with receiving ablation. In this study, we explore factors associated with receiving CA and early ablation as an interventional procedure for non-valvular AF. We use a linked administrative dataset of public and private hospital admissions in New South Wales (NSW), the most populous state in Australia.

## METHODS

### Data sources

This is a retrospective cohort study using linked administrative inpatient and mortality data from NSW, Australia. Data on AF hospitalizations were extracted from the NSW Admitted Patient Data Collection (APDC), which includes records of all inpatient separations (discharges, transfers, and deaths) from public and private hospitals in NSW. The APDC records up to 50 procedures, coded using the Australian Classification of Health Interventions (ACHI) [20] and up to 51 diagnoses, coded using the International Classification of Diseases, 10th Revision, Australian Modification (ICD-10-AM) Death records were extracted from the NSW Registry of Births, Deaths, and Marriages (RBDM) death registration file. Data linkage was performed by the NSW Ministry of Health Centre for Health Record Linkage, which reports rates of false positive and false negative links of 0.5% [21]. This study was granted ethical approval by the University of New South Wales, NSW Population and Health Services Research (HREC/18/CIPHS/56), Aboriginal Health and Medical Research Council of NSW (1503/19), and Australian Institute of Health and Welfare (EO2018/2/431) research ethics committees.

### Study cohort

The cohort included patients with a primary diagnosis of AF or atrial flutter (ICD-10-AM codes: I48, I48.0, I48.1, I48.2, I48.91) between January 2009 and October 2017, with follow-up through October 2018. The index admission was identified as the first admission with a primary diagnosis of AF between January 2009 and October 2017, with no prior admission with an AF diagnosis in any diagnosis field or prior cardioversion in a three-year look-back period. Patients under 18 years at the time of the index admission were excluded. If a single hospital stay was made up of various episodes of care, we aggregated all diagnosis and procedure codes from these episodes, and for all other fields, we kept the information from the first of these related episodes.

The primary aim of this study was to explore factors associated with receiving CA as an interventional procedure following a diagnosis of AF. As we only have access to hospital administrative data, we excluded patients whose index AF admission included an ablation procedure, under the assumption that these were patients who had been previously diagnosed in outpatient care. We excluded patients who had a diagnosis of valvular heart disease (ICD10 code I05 - rheumatic mitral valve diseases, Q23 - congenital malformations of aortic and mitral valves) or mitral valve stenosis (I34 nonrheumatic mitral valve disorders), or a replacement of mitral valve procedure (coded as ACHI codes 38488-09, 38488-02, 38488-03, 38489-02) during the look-back period or the index admission.

### Outcomes and Covariates

The primary outcome was CA for AF (ACHI codes 38290-01, 38287-02). We defined early ablation as CA administered within one year of the index AF admission. Medical history and comorbidities were determined by examining diagnosis codes (primary and otherwise) in the index AF admission and prior admissions in the three-year look-back period. Patient age and sex were obtained from the APDC record for the index AF admission. Rather than using all comorbidities from all available diagnosis codes in the cohort, we restricted selection to relevant comorbidities based on the expertise of two cardiologists (DB and RS). Prior diagnosis codes were used to calculate the AF stroke risk score (CHA_2_DS_2_-VASc [22]) when describing the characteristics of the cohort. Cardioversion was included as a covariate as it is a common procedure for AF.

Health system factors were included to assess associations between sociodemographic and health resource utilization factors with CA: year of index admission, patient payment status, and socio-economic status (SES) of their area of residence. Australia has a mixed public and private health system, and patients with private health insurance have access to shorter waiting times and greater choices in treating practitioners. Patient payment status was measured on the index admission and was categorized as public or private patients. Patients with a status of workers compensation, veteran affairs, and defense force were included with private patients.

We used patient’s Statistical Local Area of residence (Australian Statistical Geography Classification 2011 Boundaries) to measure SES using publicly available socio-economic indices of deprivation (the Index of Relative Socio-economic Advantage and Disadvantage) [23]. This index was stratified into quintiles (with quintile 1 indicating lowest relative SES and quintile 5 indicating highest SES).

### Statistical analyses

We used Cox proportional hazard models for determining the risk factors associated with receiving catheter ablation (dependent variable). The time-to-event of catheter ablation was determined by calculating the number of months from the index admission until the earliest event among 1) the first occurrence of ablation, 2) the end of the time-at-risk window (31 October 2018), and 3) death. Patients with an index admission after 31 October 2017 were excluded to ensure all patients were followed for at least one year. Among patients who received an ablation, the risk factors associated with receiving early ablation (within the first year of index AF admission) vs late ablation was determined using logistic regression. The same covariates and adjustment strategy were used for both analyses. The base model included age and sex. The first adjusted model included clinical factors: cardioversion, congestive heart failure, hypertension, diabetes, stroke, myocardial infarction (MI), coronary artery disease, and major bleeding events. The second adjusted model included health system factors: private patient status (public patient status as reference), socio-economic index of advantage and disadvantage (quintile modeled as an ordinal variable), and year of index admission. The third adjusted model included both clinical and health system factors. The models’ goodness-of-fit was evaluated using the concordance index (for the Cox models) and the area under the receiver operating characteristic curve (AUC, for the logistic regression models) under 10-fold cross-validation. Univariate survival analysis using Kaplan-Meier was estimated using the entire dataset to calculate the probability of receiving ablation vs no ablation. For readability purposes (CA is a positive event), we use the wording “likelihood” when describing the hazard ratio results. We present odds ratios for the logistic regression results.

## RESULTS

### Baseline characteristics

Table 1 shows the baseline characteristics of the entire cohort at the first AF admission (N=46,764). The ratio of male to female patients with a diagnosis of AF was almost equal (51.8% males), but the ratio of males to females receiving ablation was 2:1 (64.1% males). The patients who received ablation were younger (median age 61) compared to those who did not receive ablation (median age 74) and were more likely to have been private patients in their index admission (69.2% private patients, compared to 45% in those who did not receive ablation). Those who received ablation had lower CHA_2_DS_2_-VASc risk scores, lower comorbidities in general, and more likely to have had a cardioversion. Patients from the two upper-income quartiles received more ablations.

**Table 1.**
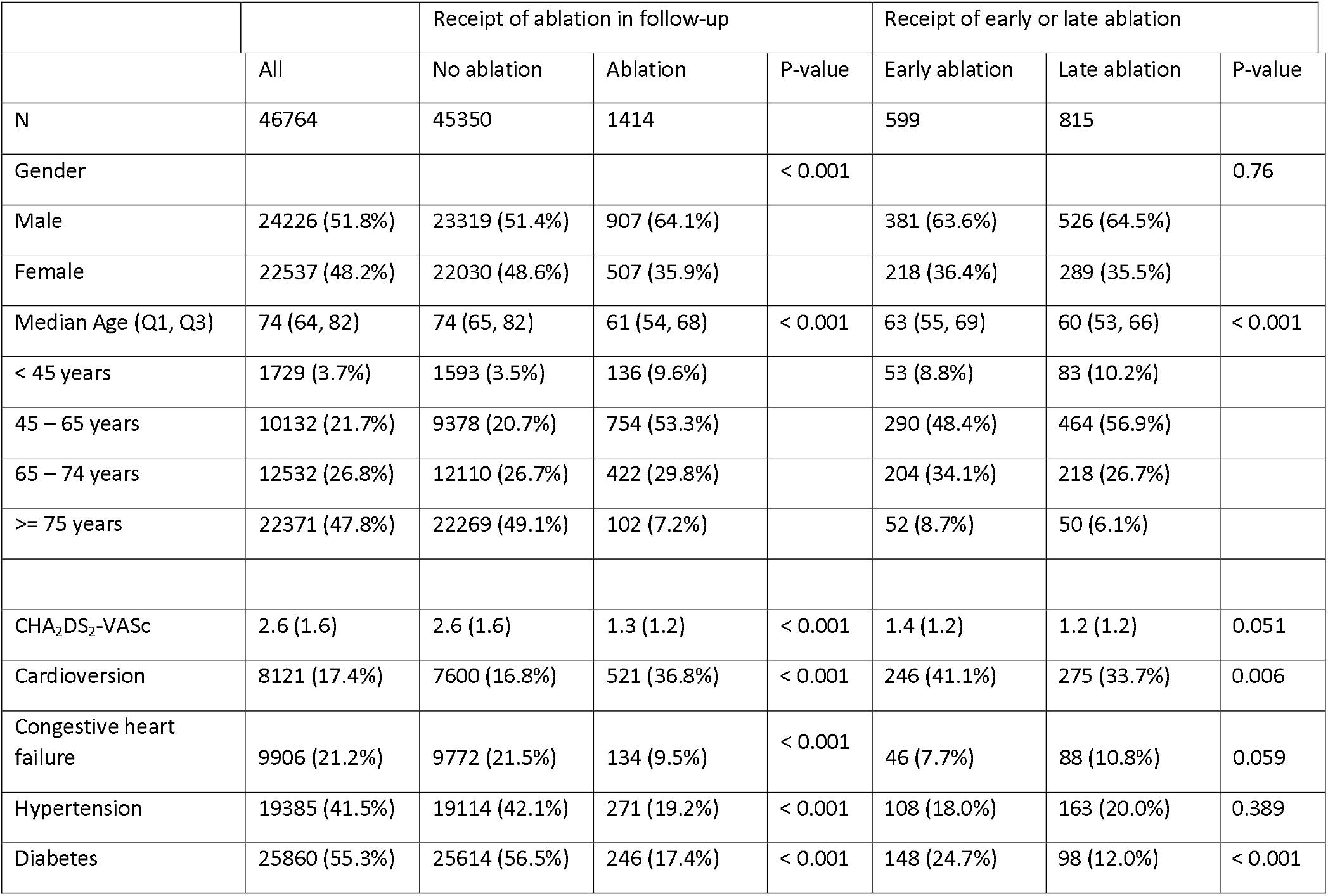
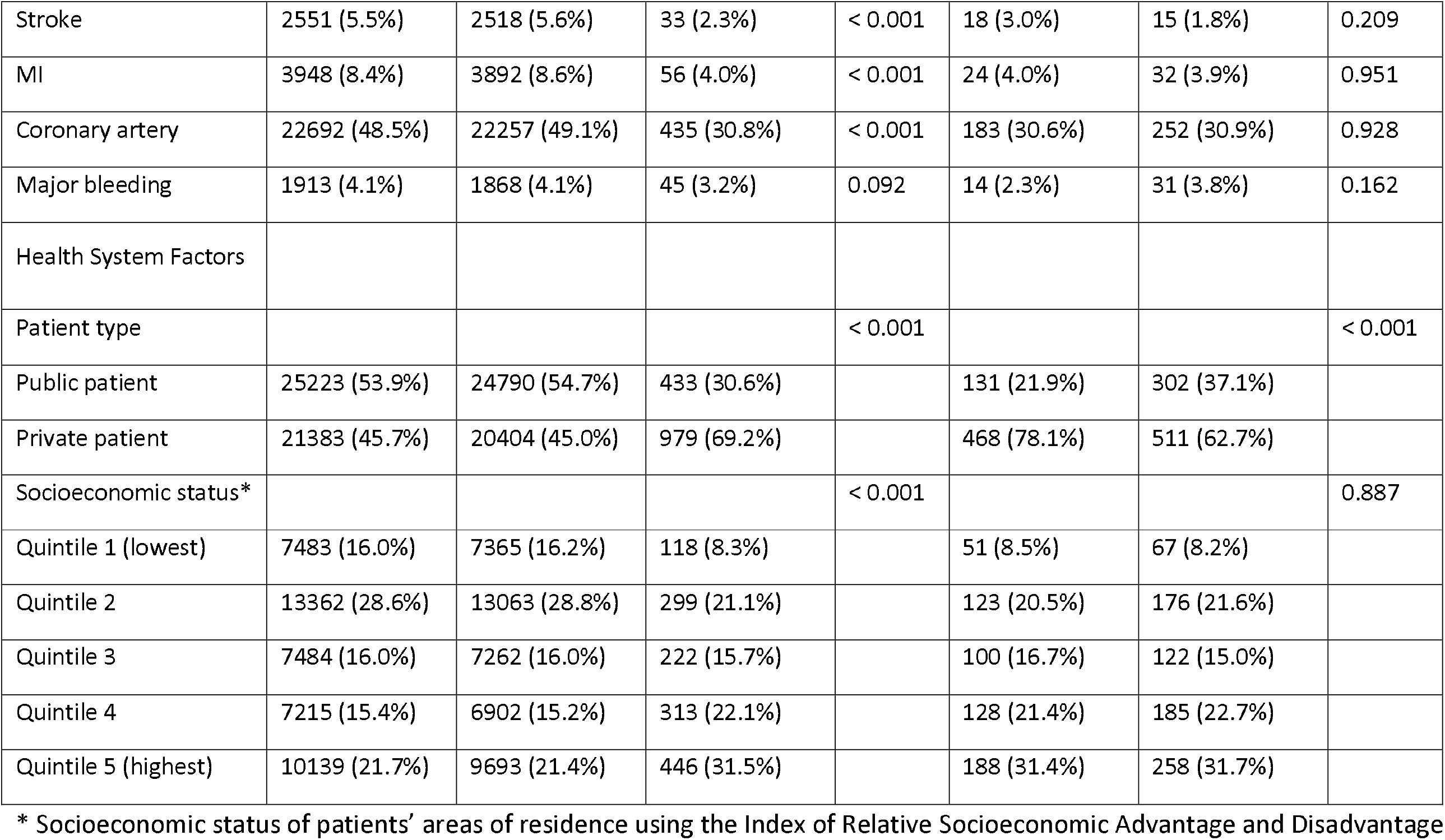
Summary of baseline characteristics of patients at time of index atrial fibrillation (AF) admission to a public or private hospital in New South Wales, Australia. Characteristics are compared for all participants and for those who receive vs do not receive ablation using all available follow-up. From the patients who receive ablation, baseline characteristics are presented for those who receive early ablation vs late ablation.

|  |  | Receipt of ablation in follow-up |  |  | Receipt of early or late ablation |  |  |
| --- | --- | --- | --- | --- | --- | --- | --- |
|  | All | No ablation | Ablation | P-value | Early ablation | Late ablation | P-value |
| N | 46764 | 45350 | 1414 |  | 599 | 815 |  |
| Gender |  |  |  | < 0.001 |  |  | 0.76 |
| Male | 24226 (51.8%) | 23319 (51.4%) | 907 (64.1%) |  | 381 (63.6%) | 526 (64.5%) |  |
| Female | 22537 (48.2%) | 22030 (48.6%) | 507 (35.9%) |  | 218 (36.4%) | 289 (35.5%) |  |
| Median Age (Q1, Q3) | 74 (64, 82) | 74 (65, 82) | 61 (54, 68) | < 0.001 | 63 (55, 69) | 60 (53, 66) | < 0.001 |
| < 45 years | 1729 (3.7%) | 1593 (3.5%) | 136 (9.6%) |  | 53 (8.8%) | 83 (10.2%) |  |
| 45 – 65 years | 10132 (21.7%) | 9378 (20.7%) | 754 (53.3%) |  | 290 (48.4%) | 464 (56.9%) |  |
| 65 – 74 years | 12532 (26.8%) | 12110 (26.7%) | 422 (29.8%) |  | 204 (34.1%) | 218 (26.7%) |  |
| >= 75 years | 22371 (47.8%) | 22269 (49.1%) | 102 (7.2%) |  | 52 (8.7%) | 50 (6.1%) |  |
| CHA<sub>2</sub>DS<sub>2</sub>-VASc | 2.6 (1.6) | 2.6 (1.6) | 1.3 (1.2) | < 0.001 | 1.4 (1.2) | 1.2 (1.2) | 0.051 |
| Cardioversion | 8121 (17.4%) | 7600 (16.8%) | 521 (36.8%) | < 0.001 | 246 (41.1%) | 275 (33.7%) | 0.006 |
| Congestive heart failure | 9906 (21.2%) | 9772 (21.5%) | 134 (9.5%) | < 0.001 | 46 (7.7%) | 88 (10.8%) | 0.059 |
| Hypertension | 19385 (41.5%) | 19114 (42.1%) | 271 (19.2%) | < 0.001 | 108 (18.0%) | 163 (20.0%) | 0.389 |
| Diabetes | 25860 (55.3%) | 25614 (56.5%) | 246 (17.4%) | < 0.001 | 148 (24.7%) | 98 (12.0%) | < 0.001 |
| Stroke | 2551 (5.5%) | 2518 (5.6%) | 33 (2.3%) | < 0.001 | 18 (3.0%) | 15 (1.8%) | 0.209 |
| MI | 3948 (8.4%) | 3892 (8.6%) | 56 (4.0%) | < 0.001 | 24 (4.0%) | 32 (3.9%) | 0.951 |
| Coronary artery | 22692 (48.5%) | 22257 (49.1%) | 435 (30.8%) | < 0.001 | 183 (30.6%) | 252 (30.9%) | 0.928 |
| Major bleeding | 1913 (4.1%) | 1868 (4.1%) | 45 (3.2%) | 0.092 | 14 (2.3%) | 31 (3.8%) | 0.162 |
| Health System Factors |  |  |  |  |  |  |  |
| Patient type |  |  |  | < 0.001 |  |  | < 0.001 |
| Public patient | 25223 (53.9%) | 24790 (54.7%) | 433 (30.6%) |  | 131 (21.9%) | 302 (37.1%) |  |
| Private patient | 21383 (45.7%) | 20404 (45.0%) | 979 (69.2%) |  | 468 (78.1%) | 511 (62.7%) |  |
| Socioeconomic status* |  |  |  | < 0.001 |  |  | 0.887 |
| Quintile 1 (lowest) | 7483 (16.0%) | 7365 (16.2%) | 118 (8.3%) |  | 51 (8.5%) | 67 (8.2%) |  |
| Quintile 2 | 13362 (28.6%) | 13063 (28.8%) | 299 (21.1%) |  | 123 (20.5%) | 176 (21.6%) |  |
| Quintile 3 | 7484 (16.0%) | 7262 (16.0%) | 222 (15.7%) |  | 100 (16.7%) | 122 (15.0%) |  |
| Quintile 4 | 7215 (15.4%) | 6902 (15.2%) | 313 (22.1%) |  | 128 (21.4%) | 185 (22.7%) |  |
| Quintile 5 (highest) | 10139 (21.7%) | 9693 (21.4%) | 446 (31.5%) |  | 188 (31.4%) | 258 (31.7%) |  |

\* Socioeconomic status of patients' areas of residence using the Index of Relative Socioeconomic Advantage and Disadvantage

Amongst patients that received ablation (see Appendix for baseline characteristics stratified by gender), females were older (median age 64) compared to men (median age 61), had higher CHA_2_DS_2_-VASc risk scores (mean score 2.0 vs 0.9), had lower rates of comorbidities (except diabetes, 21.9% of females vs 14.9% of males), were less likely to have had a cardioversion (29.6% vs 40.9%) or to have been private patients in their index admission (63.7% vs 72.3%). Men that received ablation had higher rates in the highest income quartile, whereas females had higher rates in the four lower-income quantiles.

Patients who received early ablation were slightly older (median age 63 vs median age 60), had higher rates of cardioversion and diabetes, and were more likely to have been private patients in their index admission, than patients who received a late ablation.

### Characteristics Associated with Ablation

Figure 1 shows the Kaplan Meier survival curve representing the cumulative probability of receiving ablation after index admission, with about 5% of patients receiving CA and a quarter of them occurring within the first year. The hazard ratios (and corresponding p-values) from the time-to-ablation models are shown in Table 2.

**Figure 1.**
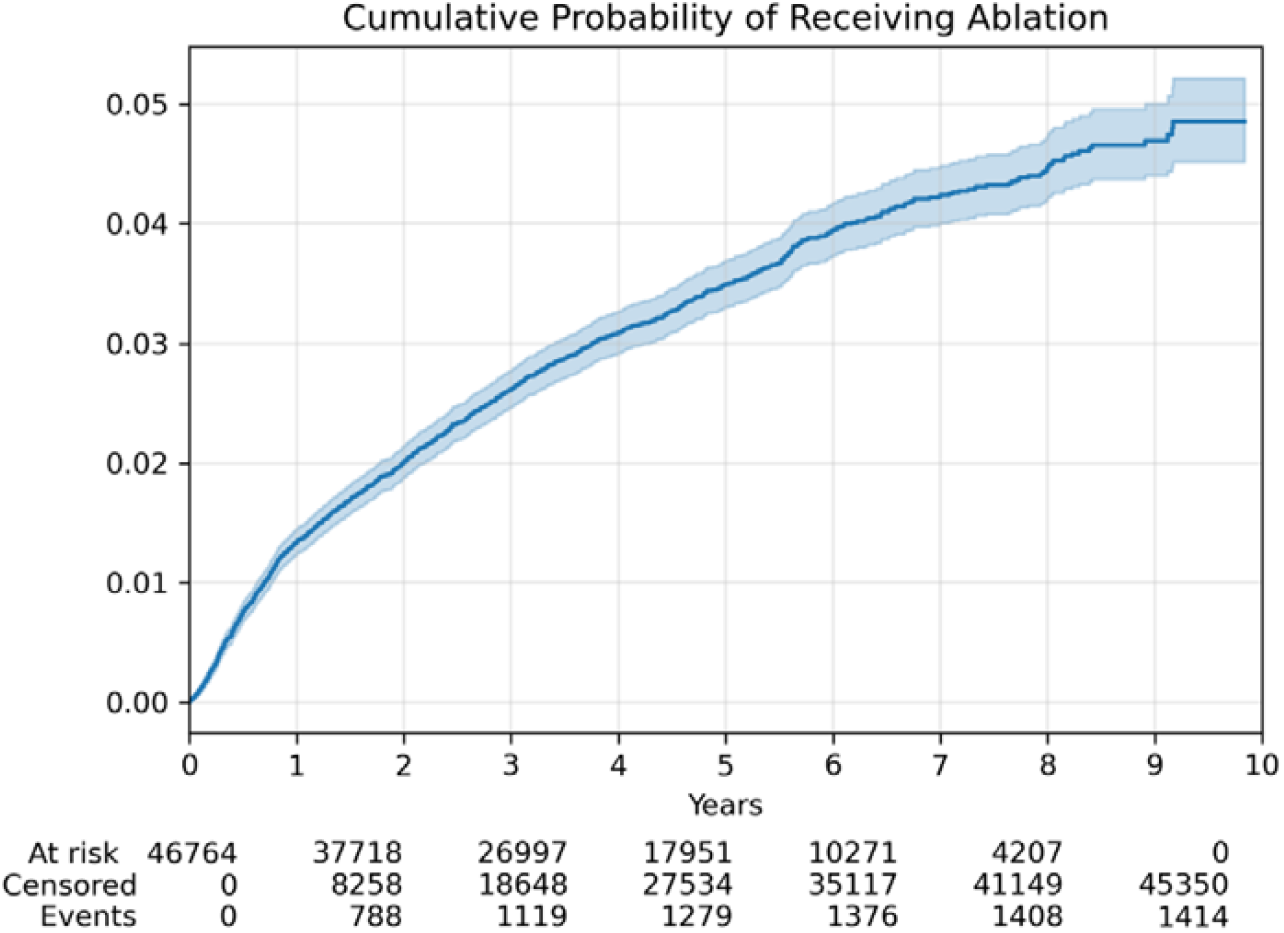
Kaplan Meier survival curve showing the cumulative probability of receiving catheter ablation after index AF admission, with about 5% of patients receiving catheter ablation, a quarter of ablations occurring within the first year and half of the ablations occurring within the first three years.

**Table 2.**
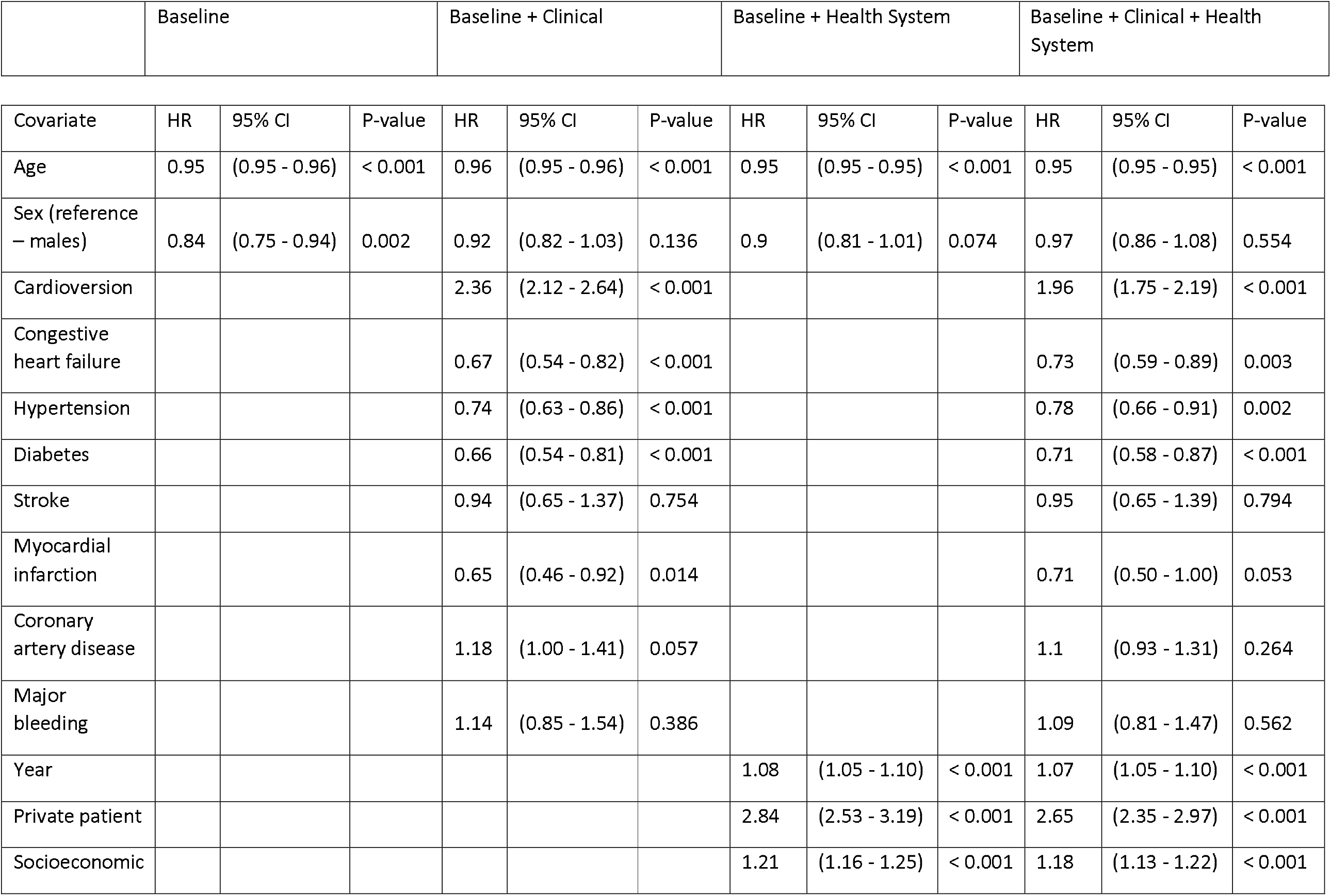
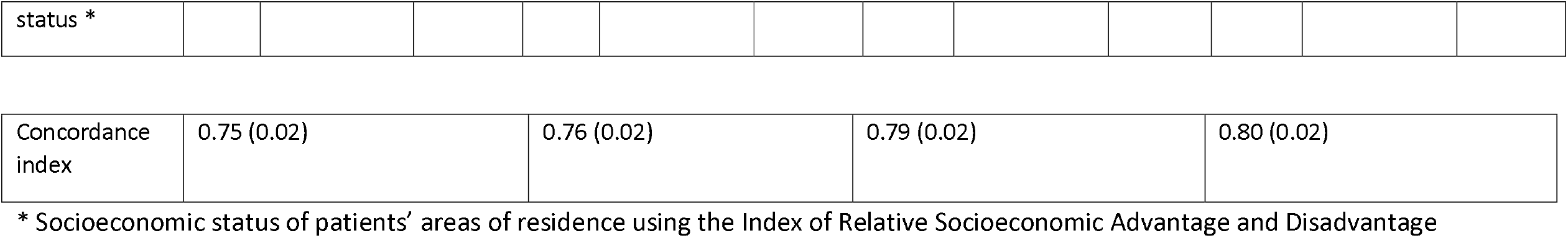
Hazard ratios from Cox regression to determine risk factors associated with receiving ablation.

|  | Baseline |  |  | Baseline + Clinical |  |  | Baseline + Health System |  |  | Baseline + Clinical + Health System |  |  |
| --- | --- | --- | --- | --- | --- | --- | --- | --- | --- | --- | --- | --- |
| Covariate | HR | 95% CI | P-value | HR | 95% CI | P-value | HR | 95% CI | P-value | HR | 95% CI | P-value |
| Age | 0.95 | (0.95 - 0.96) | < 0.001 | 0.96 | (0.95 - 0.96) | < 0.001 | 0.95 | (0.95 - 0.95) | < 0.001 | 0.95 | (0.95 - 0.95) | < 0.001 |
| Sex (reference – males) | 0.84 | (0.75 - 0.94) | 0.002 | 0.92 | (0.82 - 1.03) | 0.136 | 0.9 | (0.81 - 1.01) | 0.074 | 0.97 | (0.86 - 1.08) | 0.554 |
| Cardioversion |  |  |  | 2.36 | (2.12 - 2.64) | < 0.001 |  |  |  | 1.96 | (1.75 - 2.19) | < 0.001 |
| Congestive heart failure |  |  |  | 0.67 | (0.54 - 0.82) | < 0.001 |  |  |  | 0.73 | (0.59 - 0.89) | 0.003 |
| Hypertension |  |  |  | 0.74 | (0.63 - 0.86) | < 0.001 |  |  |  | 0.78 | (0.66 - 0.91) | 0.002 |
| Diabetes |  |  |  | 0.66 | (0.54 - 0.81) | < 0.001 |  |  |  | 0.71 | (0.58 - 0.87) | < 0.001 |
| Stroke |  |  |  | 0.94 | (0.65 - 1.37) | 0.754 |  |  |  | 0.95 | (0.65 - 1.39) | 0.794 |
| Myocardial infarction |  |  |  | 0.65 | (0.46 - 0.92) | 0.014 |  |  |  | 0.71 | (0.50 - 1.00) | 0.053 |
| Coronary artery disease |  |  |  | 1.18 | (1.00 - 1.41) | 0.057 |  |  |  | 1.1 | (0.93 - 1.31) | 0.264 |
| Major bleeding |  |  |  | 1.14 | (0.85 - 1.54) | 0.386 |  |  |  | 1.09 | (0.81 - 1.47) | 0.562 |
| Year |  |  |  |  |  |  | 1.08 | (1.05 - 1.10) | < 0.001 | 1.07 | (1.05 - 1.10) | < 0.001 |
| Private patient |  |  |  |  |  |  | 2.84 | (2.53 - 3.19) | < 0.001 | 2.65 | (2.35 - 2.97) | < 0.001 |
| Socioeconomic |  |  |  |  |  |  | 1.21 | (1.16 - 1.25) | < 0.001 | 1.18 | (1.13 - 1.22) | < 0.001 |
| status * |  |  |  |  |  |  |  |  |  |  |  |  |

|  |  |  |  |  |
| --- | --- | --- | --- | --- |
| Concordance index | 0.75 (0.02) | 0.76 (0.02) | 0.79 (0.02) | 0.80 (0.02) |

\* Socioeconomic status of patients' areas of residence using the Index of Relative Socioeconomic Advantage and Disadvantage

Older age was associated with a lower likelihood of receiving CA (likelihood decreasing for each additional year of age), even after adjusting for clinical and health system factors. Female sex was associated with a lower likelihood of receiving CA in the base model, but not after adjusting for clinical and health system factors. Cardioversion during index admission was associated with a higher likelihood of receiving CA, even after adjusting for health system factors. A history of congestive heart failure, hypertension, diabetes, and MI were associated with a lower likelihood of receiving CA, even after adjusting for health system factors. A history of stroke, coronary artery disease, and major bleeding events were not associated with a lower or higher likelihood of receiving CA. All health system factors were associated with a higher likelihood of receiving CA even after adjustment for clinical factors, including non-public payment (private patient), as well as increasing SES of the area of residence. For the goodness-of-fit, the model with health system covariates had a higher concordance index compared to the model with only clinical factors (Table 2).

### Characteristics Associated with Early Ablation

Table 3 displays the odds ratios associated with receiving early ablation. Cardioversion during index admission and history of diabetes were associated with a higher likelihood of early ablation, even after adjusting for health system covariates. Private patient status was associated with a higher likelihood of early ablation even after adjusting for clinical covariates. The model with the health system factors had a higher AUC than the model with clinical factors.

**Table 3.**
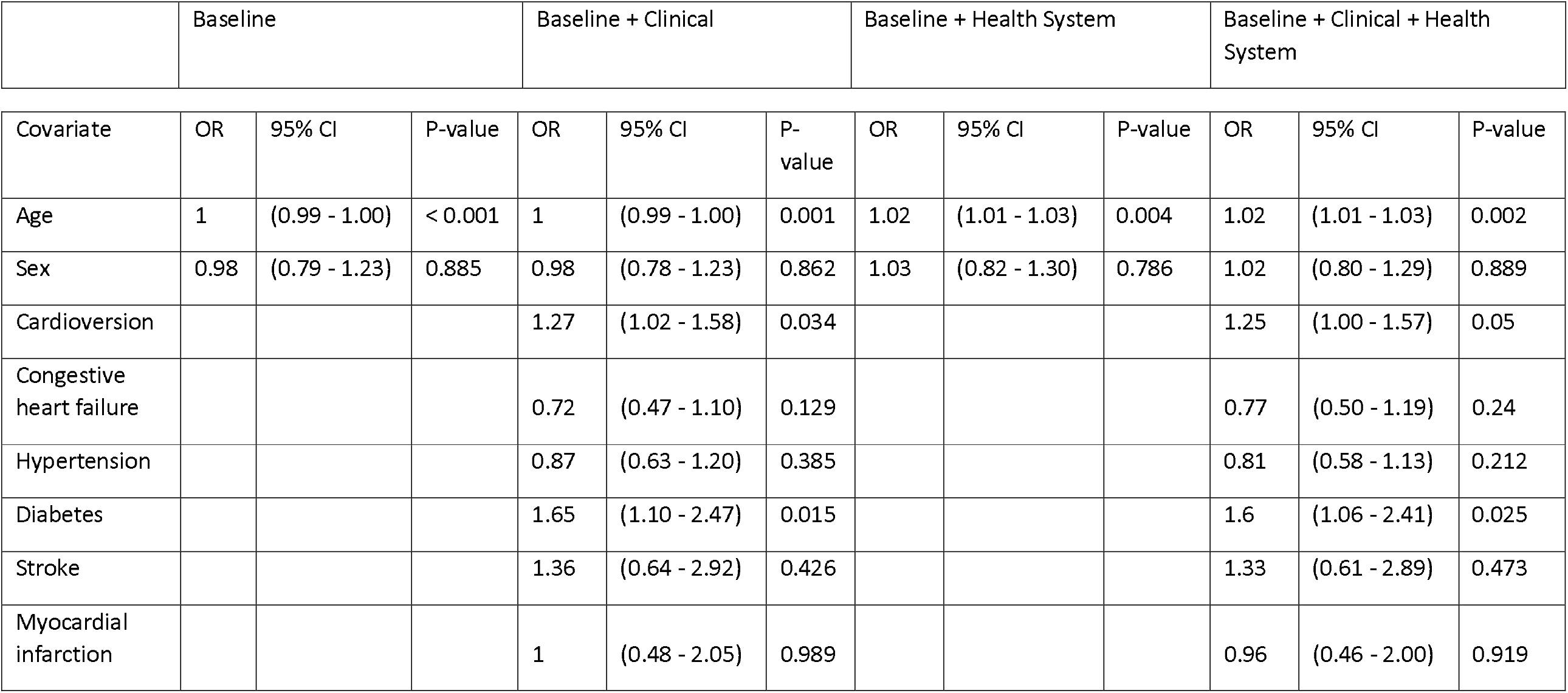
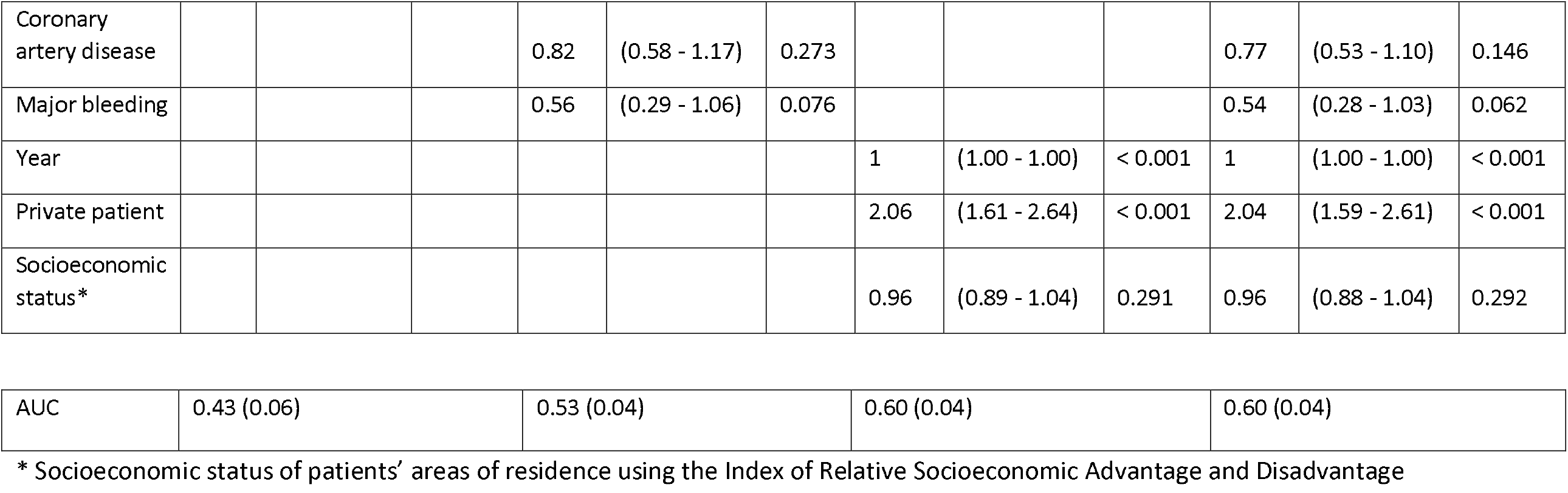
Odds ratios from logistic regression to determine risk factors associated with receiving early ablation.

|  | Baseline |  |  | Baseline + Clinical |  |  | Baseline + Health System |  |  | Baseline + Clinical + Health System |  |  |
| --- | --- | --- | --- | --- | --- | --- | --- | --- | --- | --- | --- | --- |
| Covariate | OR | 95% CI | P-value | OR | 95% CI | P-value | OR | 95% CI | P-value | OR | 95% CI | P-value |
| Age | 1 | (0.99 - 1.00) | < 0.001 | 1 | (0.99 - 1.00) | 0.001 | 1.02 | (1.01 - 1.03) | 0.004 | 1.02 | (1.01 - 1.03) | 0.002 |
| Sex | 0.98 | (0.79 - 1.23) | 0.885 | 0.98 | (0.78 - 1.23) | 0.862 | 1.03 | (0.82 - 1.30) | 0.786 | 1.02 | (0.80 - 1.29) | 0.889 |
| Cardioversion |  |  |  | 1.27 | (1.02 - 1.58) | 0.034 |  |  |  | 1.25 | (1.00 - 1.57) | 0.05 |
| Congestive heart failure |  |  |  | 0.72 | (0.47 - 1.10) | 0.129 |  |  |  | 0.77 | (0.50 - 1.19) | 0.24 |
| Hypertension |  |  |  | 0.87 | (0.63 - 1.20) | 0.385 |  |  |  | 0.81 | (0.58 - 1.13) | 0.212 |
| Diabetes |  |  |  | 1.65 | (1.10 - 2.47) | 0.015 |  |  |  | 1.6 | (1.06 - 2.41) | 0.025 |
| Stroke |  |  |  | 1.36 | (0.64 - 2.92) | 0.426 |  |  |  | 1.33 | (0.61 - 2.89) | 0.473 |
| Myocardial infarction |  |  |  | 1 | (0.48 - 2.05) | 0.989 |  |  |  | 0.96 | (0.46 - 2.00) | 0.919 |
| Coronary artery disease |  |  |  | 0.82 | (0.58 - 1.17) | 0.273 |  |  |  | 0.77 | (0.53 - 1.10) | 0.146 |
| Major bleeding |  |  |  | 0.56 | (0.29 - 1.06) | 0.076 |  |  |  | 0.54 | (0.28 - 1.03) | 0.062 |
| Year |  |  |  |  |  |  | 1 | (1.00 - 1.00) | < 0.001 | 1 | (1.00 - 1.00) | < 0.001 |
| Private patient |  |  |  |  |  |  | 2.06 | (1.61 - 2.64) | < 0.001 | 2.04 | (1.59 - 2.61) | < 0.001 |
| Socioeconomic status* |  |  |  |  |  |  | 0.96 | (0.89 - 1.04) | 0.291 | 0.96 | (0.88 - 1.04) | 0.292 |

|  |  |  |  |  |
| --- | --- | --- | --- | --- |
| AUC | 0.43 (0.06) | 0.53 (0.04) | 0.60 (0.04) | 0.60 (0.04) |

\* Socioeconomic status of patients' areas of residence using the Index of Relative Socioeconomic Advantage and Disadvantage

## DISCUSSION

### Main findings

Age was associated with a lower likelihood of receiving CA (likelihood decreasing for each additional year). Patients with a cardioversion during index admission were more likely to receive CA, consistent with prior work showing an association between the risk of ablation and the number of performed cardioversions [19]. This likely reflects a desire for a “rhythm control” strategy in these patients, based on clinical factors. Women in our cohort, regardless of receipt of ablation, received fewer cardioversions than men. Patients with a medical history of congestive heart failure, hypertension, and diabetes being less likely to receive CA is consistent with other studies [10], [17] and suggests that ablation is performed on relatively healthier patients (fewer comorbidities), who are at an earlier stage in the disease process and more likely to have a successful procedure. Health system factors associated with an increased likelihood of receiving CA included the year of index AF admission (risk increasing with year), private patients, and patients from areas of higher socioeconomic status. Patients with a cardioversion during index admission, a history of diabetes, and who were private patients (at their index admission) had higher odds of receiving early ablation.

Of all model adjustments for determining factors associated with CA and early ablation, the models with health system covariates best explained the data. This is alarming, as it shows health system factors being stronger predictors than clinical factors.

### Impact of Health System Factors on Receiving Ablation

Our findings suggest the increased use of ablation as a procedural intervention for AF patients over time, consistent with studies in Australia [13] and the US [17]. The strong association of private patient status with ablation suggests that patients who enter the health system for their care of AF through the private system (or even through other forms of subsidized care) have faster access to ablation. These findings most likely reflect that private patients in Australia have increased access to elective procedures through their health insurance, including shorter waiting times and choice of treating practitioner. Our results also highlight that patients living in more advantaged areas are more likely to receive ablation even after accounting for insurance status, suggesting there may also be inequities in access to the procedure – either through capacity for affording out-of-pocket costs, or in the geographic location of services. The association between private patient status and early ablation highlights that public patients may first undergo alternative or lower-cost care, with ablation performed at a later stage. Whether this delayed access is resulting in different outcomes for public and private patients requires further investigation.

A study with a cohort of under 65 years of age in the US found insurance type to be a predictor of ablation [17] and a study using data from Swedish health registries showed that university education and income in the highest quartile were factors associated with undergoing ablation [18]. More recently, a study comparing surgical utilization in NSW and other countries showed that residents of lower-income neighborhoods had lower rates of surgery compared to residents of higher-income neighborhoods [24]. Our work similarly highlights disparities in receiving CA associated with patient status and the neighborhood index of social advantage and disadvantage, raising concerns on the equitable delivery of CA in Australia. However, administrative data is unlikely to capture the complexity of the risk-benefit analysis in individual patients, and there may be further socio-demographic and health system factors that influence clinical recommendation and patient utilization of ablation procedures.

### Limitations

This study used linked administrative records from hospitalizations in NSW, Australia. These do not capture information about the use of antiarrhythmic medications and anticoagulants, family history, medical history, social and lifestyle factors, and any diagnosis or procedure from outpatient care. As such, the date of index AF admission is a proxy for date of AF diagnosis. Our analysis relied solely on coded hospital diagnoses, which are recorded only for conditions that significantly affect patient management during an episode of care. As such, our use of diagnoses as a proxy for incident or prevalent disease, the inferring of date of onset of AF, and the quality of hospital diagnosis coding varying widely (accuracy 51-98% across 32 studies) [25] are potential sources of bias in our analysis. However, this source of bias is lessened by our use of a large dataset, hospitalizations coming from public and private hospitals across the state of NSW. Finally, our survival model did not take into consideration competing interventions (open ablation) or death as a competing risk, which may have produced a slight overestimation in the results.

## CONCLUSION

In a cohort of patients with non-valvular AF, using linked administrative data from Australia’s most populous state (NSW), this study showed potential disparities in the likelihood of receiving ablation and early ablation between public and private patients. Clinicians and policymakers should review existing policies to ensure effective procedures for AF are available to the whole population.

## Data Availability

Not applicable

## Author contributions

All authors contributed to the conception and design of the study. JCQ performed the data analysis and wrote the first draft. All authors contributed to critical revisions of the manuscript. All authors approved the final draft.

## Acknowledgements

The NSW Centre for Health Record Linkage (CHeReL) for linking the datasets NSW Admitted Patients Data Collection and the NSW Registry of Births, Deaths, and Marriages. The NSW Ministry of Health for providing access to population health data. Dr. Oscar Perez-Concha, Dr. Malcolm Gillies, and Dr. Mark Hanly for statistical feedback. Dr. Benjamin Hsu for providing access to the datasets. Dr. Juliana Costa for feedback on cohort generation and coding of transfer episodes.

## Funding

This work was supported by National Health and Medical Research Council (NHMRC), project grant No. APP1184304. Creation of the linked dataset was funded by a NHMRC Project Grant No. 1147430. MOF is supported by an NHMRC Early Career Fellowship No. 1139133.

## Competing interests

The authors declare no competing interests.

## Appendix

**Table 4.**
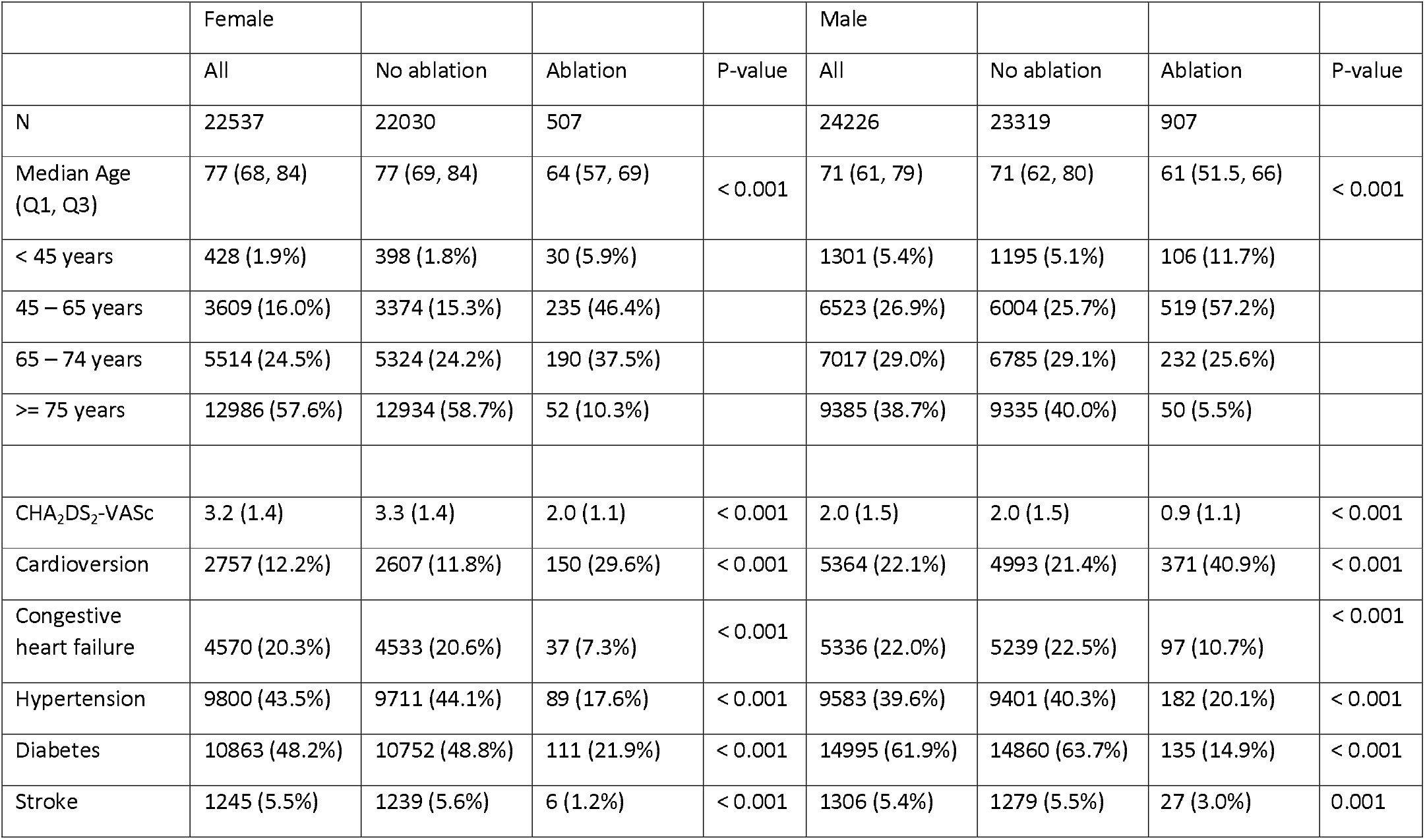
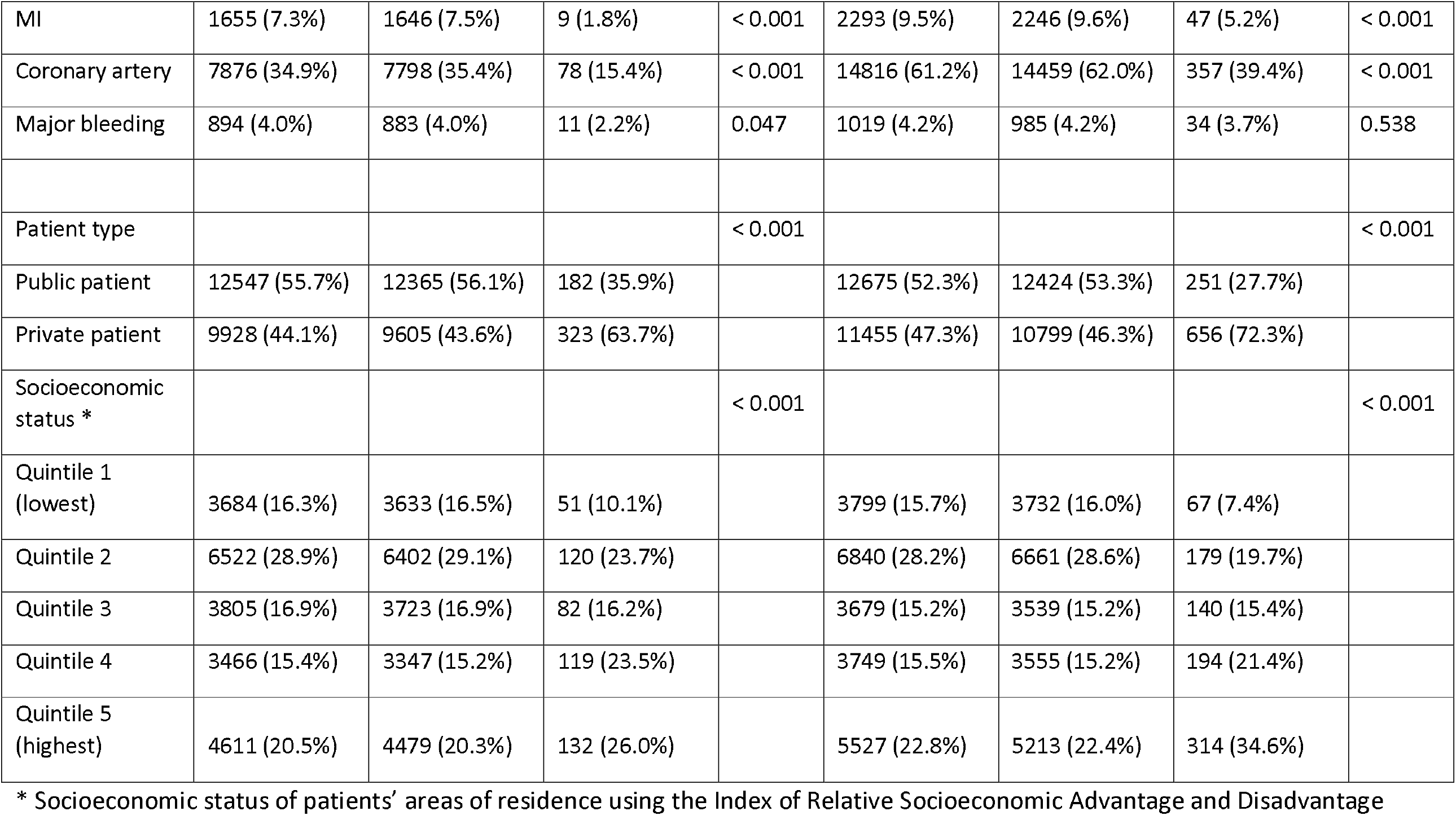
Summary of baseline characteristics of patients (stratified by gender) at time of index atrial fibrillation (AF) admission to a public or private hospital in New South Wales, Australia. Characteristics are compared for all participants and for those who receive vs do not receive ablation.

## Notes

### Competing Interest Statement

The authors have declared no competing interest.

### Author Declarations

This study was granted ethical approval by the University of New South Wales, NSW Population and Health Services Research (HREC/18/CIPHS/56), Aboriginal Health and Medical Research Council of NSW (1503/19), and Australian Institute of Health and Welfare (EO2018/2/431) research ethics committees.

